# Relationship between Average Daily Temperature and Average Cumulative Daily Rate of Confirmed Cases of COVID-19

**DOI:** 10.1101/2020.04.10.20059337

**Authors:** Behzad Pirouz, Amirsina Golmohammadi, Hasti SaeidpourMasouleh, Galileo Violini, Behrouz Pirouz

## Abstract

**AIMS:** The main purpose of this study is to investigate the correlation between the average daily temperature and the rate of coronavirus epidemic growth in the infected regions.

**BACKGROUND:** The rapid outbreak of the new Coronavirus (COVID-19) pandemic and the spread of the virus worldwide, especially in the Northern Hemisphere, have prompted various investigations about the impact of environmental factors on the rate of development of this epidemic. Different studies have called attention to various parameters that may have influenced the spread of the virus, and in particular, the impact of climatic parameters has been emphasized.

**OBJECTIVE:** The main hypothesis object of our research is that between regions exhibiting a significant difference in the mean daily temperature, a significant difference is also observed in the average cumulative daily rate of confirmed cases and that this does not happen if there is no significant difference in mean daily temperature.

**METHOD:** The research hypothesis was investigated through statistical analysis. The F-test was used to test whether there is significant equality of variances for each pair of case studies, and then, by the T-Test, the existence of a significant difference was investigated. In all statistical tests, the confidence level of 95% is considered. In order to minimize the impact on the results of factors like the policy of the government or cultural differences among countries (food, exercise, weight, etc.), three case studies within five countries, namely Iran, Italy, Germany, Spain, and United States were compared separately.

**RESULT:** This statistical analysis shows that there is a correlation between the average temperature and the epidemic rate, and this is especially evident when differences in average daily temperature are significantly larger, as it happens for Bandar Abbas in Iran, Milan in Italy, Santa Cruz in Spain, and Los Angeles in the US. Besides, the analysis of the average air temperatures shows that the epidemic rates of COVID-19 were higher in the case studies with a lower average temperature. Instead, when no significant differences exist in the average daily temperature of two cities in the same country, there is no significant difference in the average cumulative daily rate of confirmed cases.

**CONCLUSION:** In all five selected countries, we found that when there is a significant difference in the daily mean temperature between two regions of a country, a significant difference also exists in the average cumulative daily rate of confirmed cases. Conversely, if there are no significant differences in the mean daily temperature of two regions in the same country, no significant difference is observed in the average cumulative daily rate of confirmed cases for these regions. In conclusion, the results of this study support the research hypothesis and confirm the effectiveness of the proposed method for analysis of the epidemic rates.

## 1. Introduction

The outbreak of infectious diseases has always been one of the most important health problems in the world. With the rapid spread of the new coronavirus (COVID-19), much attention has been paid to subjects like rate of an epidemic, transition ways, prevention methods, and remaining time of the virus in the environment [1-5]. Therefore, the behavior of the virus in relation to environmental factors is critical. The coronavirus is one of the epidemic diseases that many countries have experienced in the new type of it, COVID-19, and can become a serious challenge and affect the previous efforts on sustainable development [6-9].

It has been found that there are some delays between the detection of confirmed cases and the actual infection date due to the laboratory test, the time of the announcement of confirmed cases in media, and the time for the patients showing initial symptoms for a total usually of 3 to 5 days [10-12]. The analysis of Kampf et al. (2020) demonstrated that COVID-19 could remain up to 9 days on different materials based on temperature and humidity [13].

Cascella et al. (2020) conducted research on the new coronavirus structure. Their analysis shows that the virus belongs to the family of single-stranded RNA viruses (+ ssRNA), which has a length of about 30 kb and an envelope with spear structures and they tested the sensitivity of the virus to UV rays and temperature [14]. Pirouz et al. (2020) investigated the correlations between environmental factors and the number of positive cases of COVID-19 by the use of artificial intelligence (AI) techniques. The results of this study put evidence on the role of urban and weather conditions on the epidemic rate [15]. Chen et al. (2020) developed a time-dependent mathematical model for the prediction of the total number of positive cases [16].

Lytle and Sagripanti (2005) studied the impact of UV rays, and their results determined that it can affect the structure of coronaviruses because UV rays could damage the nucleic acids of viral RNA that is vital for the survival of the virus [17]. Pirouz et al., (2020) developed an assessment method for investigating the impact of climate and urban factors in confirmed cases of COVID-19 [18]. Bukhari et al., (2020), studied the epidemic of COVID-19 on tropical zones and their results show that the highest number of infection happened in temperature among 3 to 17 °C [19].

The analysis of Grant and Giovannucci (2009) shows that by increasing the UV rays, the level of vitamin D in the body increases and makes the infection rates decrease. Therefore, in areas on lower latitudes and closer to the equator line, the epidemic on COVID-19 might be different [20]. In the study of Chan et al. (2011), the effect of temperature and humidity on SARS coronavirus was analyzed, and they found that increasing temperature decreases the epidemic rate by faster disabling virus structure on contaminated surfaces [21].

Pirouz and Violini (2020), analyzed the outbreak patterns of COVID-19 by analysis of 117 countries. They suggested two mechanisms of triggering the development of the infection, namely the normal spread of an infection from one parent case and the consequence of a unique phenomenon of accumulation of contagious people [22].

Other studies in this area include Nkengasong and Mankoula, (2020), Wen et al., (2019), Gilbert et al., (2020), Chinazzi et al., (2020), and Li and Feng, (2020) [23-27].

Analysis of the previous studies demonstrates the necessity of further research about the effect of environmental factors on the COVID-19. Since some of the previous researches have shown the effect of weather conditions, especially temperature, on coronavirus viruses, the main purpose of this study is to investigate the better correlation of daily mean air temperature on the epidemic rate of the COVID-19.

## 2. Materials and Methods

In this study, we analyzed the number of positive contagions of coronavirus COVID-19, from February 29^th^ to March 19^th^. The selection of February 29^th^ is due to existence on this date of at least one positive case of COVID-19 in all case studies studied. The analysis of the weather data started from February15^th^ and ended on March 18^th^ because the impact of the weather parameter would occur in the upcoming days. Thus, the considered average daily temperature is with an interval of 1 to 14 days before the definitive cases of COVID-19.

Analysis conditions:

- In order to minimize the impact of other parameters on the results, such as the policy of each government, cultural differences between countries, health care systems, or the protocols in getting coronavirus test, comparative analysis have been done for different regions of the same country, without comparing with each other the results of different countries;
- The analysis period for average daily temperature ranges from February15^th^, 2020 to March18^th^, 2020 (33 days);
- The analysis period for confirmed cases of coronavirus COVID-19 is from February 29^th^ to March19^th^ ;
- For what concerns the cumulative daily rate of confirmed cases, the total number of confirmed cases in a region was divided into the population of that region in order to take into account the different size of the population in each region.

### 2.1. Case study

To carry out the analysis, the datasets of five countries and in each country, of three regions/provinces have been considered. The selected case studies are presented in Table 1. and the location of the three regions shown in Figures 1.

**Table 1.** The selected case studies [28, 29]

| Country | Regions/province | Population | Selected city for weather data |
| --- | --- | --- | --- |
| Iran | Mazandaran | ≈3,704,000 | Ramsar |
|  | Tehran | ≈13,270,000 | Tehran |
|  | Hormozgan | ≈1,776,000 | Bandar Abbas |
| Italy | Lombardy | 10,060,574 | Milan |
|  | Lazio | 5,879,082 | Rome |
|  | Campania | 5,801,692 | Naples |
| Germany | Berlin | 3,613,495 | Berlin |
|  | North Rhine-Westphalia | 17,912,134 | Dusseldorf |
|  | Bavaria | 12,997,204 | Munich |
| Spain | Catalonia | 7,619,494 | Barcelona |
|  | Valencia | 2,304,999 | Valencia |
|  | Canary Islands | 2,220,279 | Santa Cruz |
| United States | New York | 19,453,561 | New York |
|  | Washington | 7,614,893 | Seattle |
|  | California | 39,512,223 | Los Angeles |

**Figure 1.**
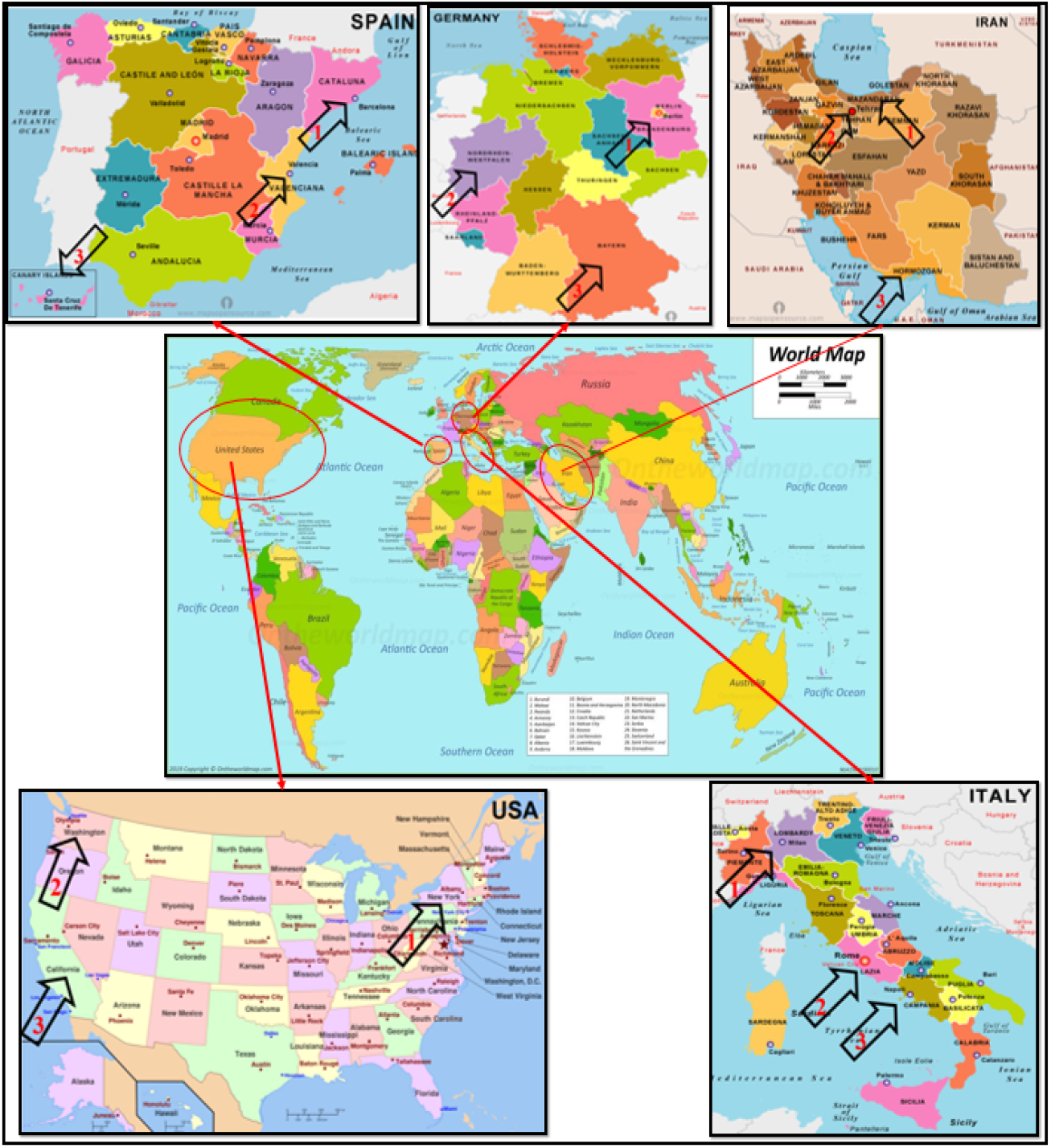
Location of the selected case studies [30, 31]

### 2.2. The statistical analysis and set-up of SPSS Model

Mathematical models have been successfully used for the analysis of many hazards [32, 33]. In complex subjects, different mathematical models can be used to solve the problem [34-36]. Statistical tests are an appropriate method for the investigation of possible correlations between different datasets [37]. Many statistical tests can be used for the analysis of significant differences between the datasets, among which F-test and Independent-Samples T-Test [38].

In order to investigate the hypothesis about a correlation between significant differences (sig.) of average daily temperatures and average cumulative daily rate of confirmed cases in two regions, the following assumptions, and statistical tests have been done:

- The variances of the two groups are equal: H0
- The variances of the two groups are not equal: H1

When using F-test, if the significance level (sig.) is less than 0.05, hypothesis H0 is rejected, and then hypothesis H1 is accepted, i.e., the variances of the datasets in the two selected regions are not equal. Conversely, if the significance level (sig.) is larger than 0.05, the hypothesis H1 is rejected, and the hypothesis H0 is accepted, i.e., the variances of the datasets in the two selected regions are equal.

Through F-test, one determines the presence or absence of differences in the variances of the two regions. Through another statistical test, the Independent-Samples T-Test, one can determine whether the difference is significant, has been used. When using Independent-Samples T-Test, a significance level (sig.) lower (higher) than 0.05, means that the difference is (is not) significant.

In our analysis, we used SPSS software. The critical factors in the SPSS Model set up are as follow:

- The datasets in different regions of the same countries were analyzed with respect to the average daily air temperature during the 33-day period from 15-Feb to 18-Mar and the cumulative daily rate of confirmed cases of COVID-19 during the 19 days from 29- Feb to 19-Mar.
- In all statistical tests, the confidence level of 95% is considered.
- First, the F-test was used to test whether there is a significant equality of variances for each pair of case studies, and then, depending on the F-test result, by the T-Test the existence of a significant difference between the data of the two cases was investigated.

## 3. Results

The average daily temperature [39, 40], and cumulative daily rate of confirmed cases [41], are shown in Figures 2 to 6.

**Figure 2.**
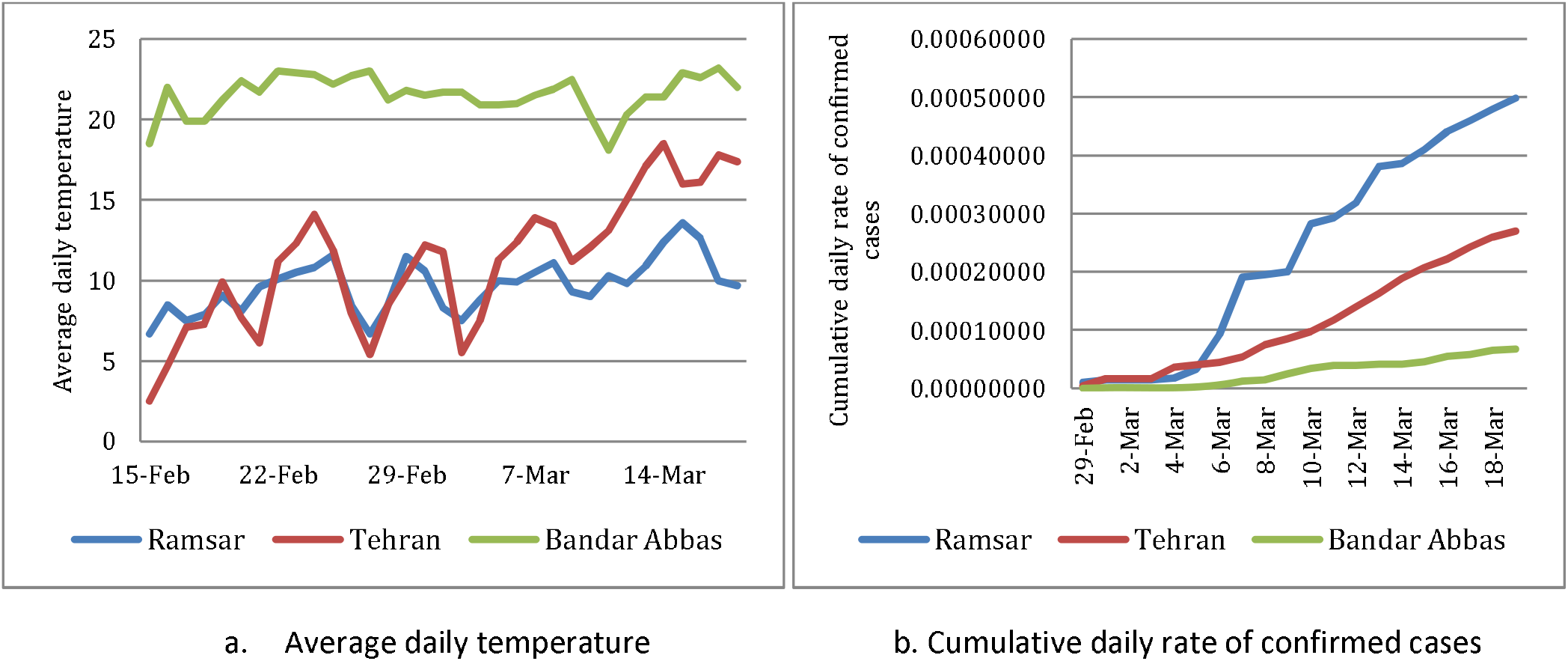
Comparison of three case studies of Iran

**Figure 3.**
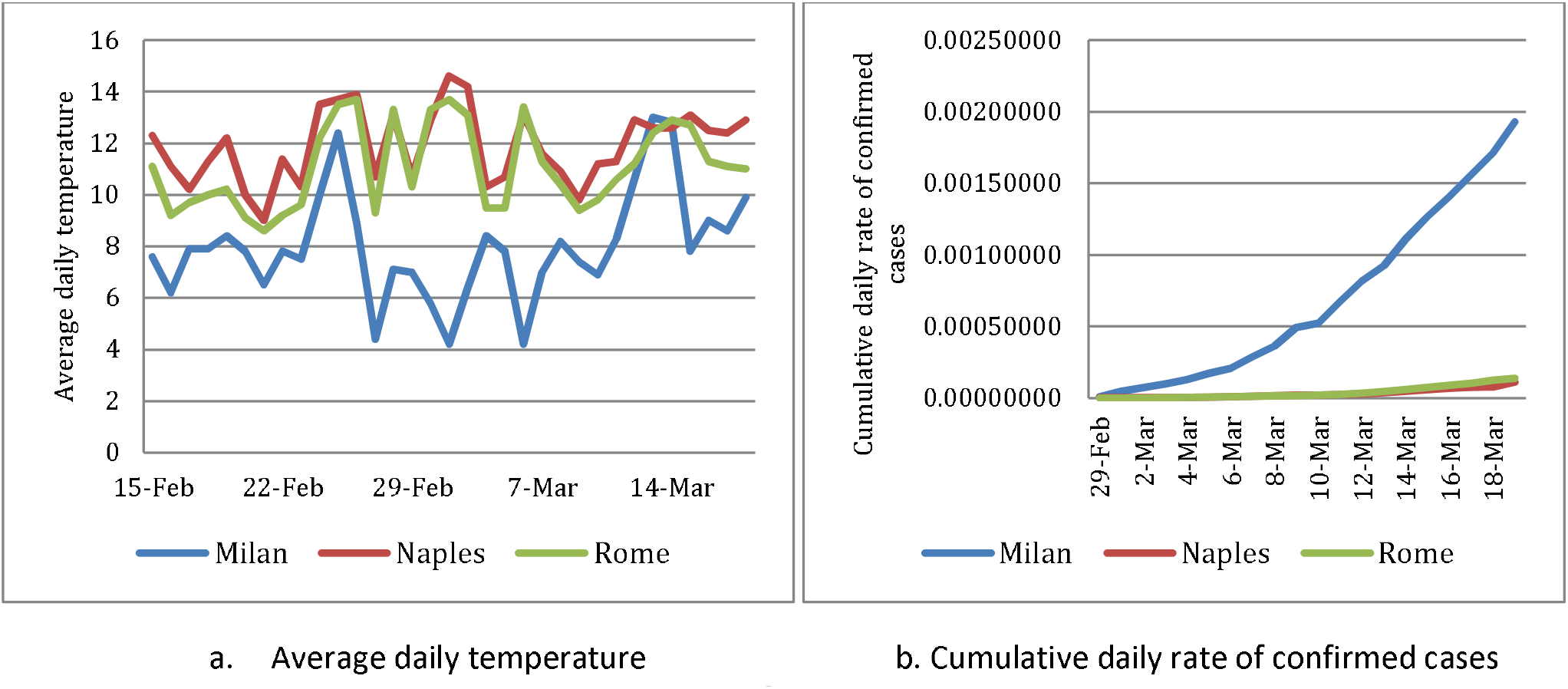
Comparison of three case studies in Italy

**Figure 4.**
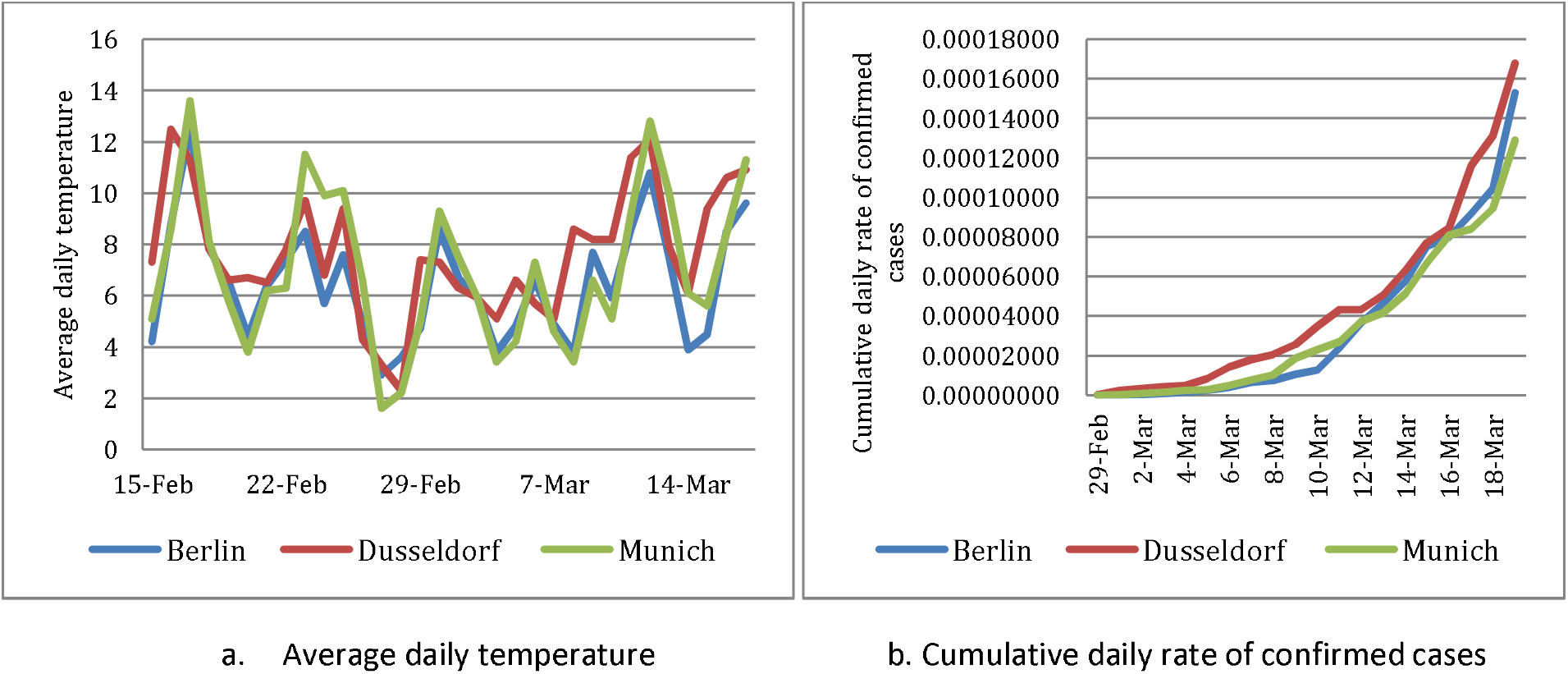
Comparison of three case studies in Germany

**Figure 5.**
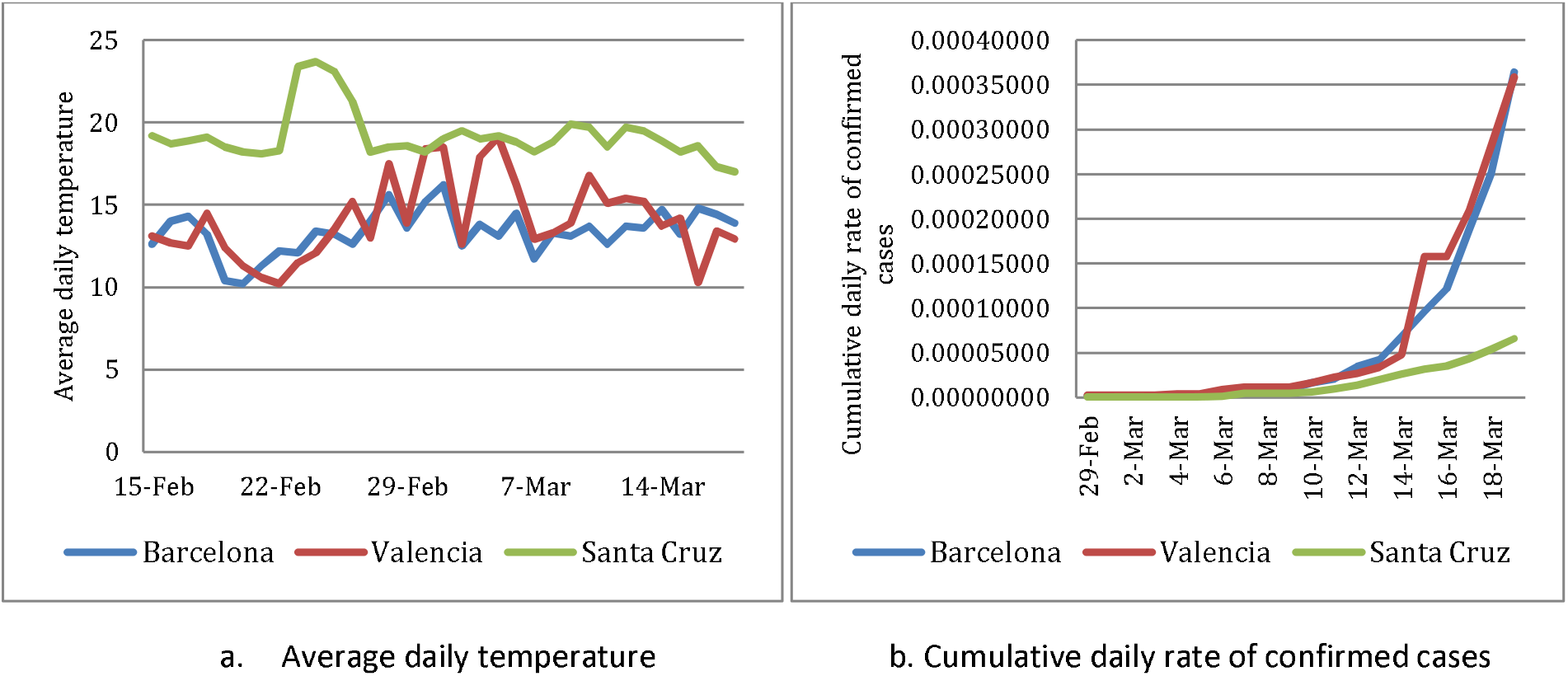
Comparison of three case studies in Spain

**Figure 6.**
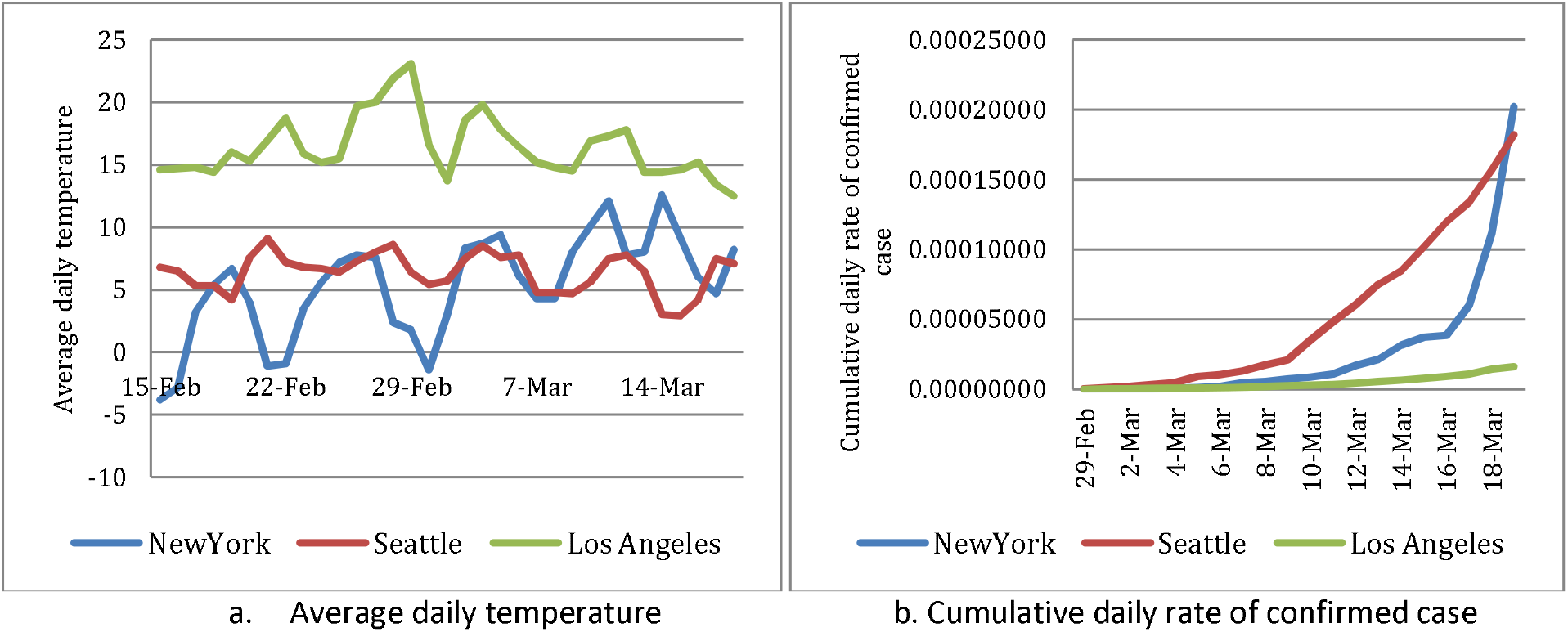
Comparison of three case studies in the United States

Correlations between the average temperatures and the cumulative daily rate of confirmed cases were found especially for towns with significant differences, with respect to the other two towns of the same country, in the average daily temperature. This was the case for Bandar Abas in Iran, Milan in Italy, Santa Cruz in Spain, and Los Angeles in the US. The average daily temperature and cumulative daily rate of confirmed cases are listed in Table 2.

**Table 2.** Average temperature and daily rate of confirmed cases

| Country | Case Study | Average Daily Temperature, °C | Average Cumulative Rate of Confirmed Cases |
| --- | --- | --- | --- |
| Iran | Ramsar | 9.69 | 0.000236 |
|  | Tehran | 11.19 | 0.000115 |
|  | Bandar Abbas | 21.54 | 0.000027 |
| Italy | Milan | 7.99 | 0.000690 |
|  | Rome | 11.08 | 0.000039 |
|  | Naples | 11.91 | 0.000031 |
| Germany | Berlin | 6.63 | 0.000036 |
|  | Dusseldorf | 7.72 | 0.000046 |
|  | Munich | 7.14 | 0.000034 |
| Spain | Barcelona | 13.35 | 0.000062 |
|  | Valencia | 14.05 | 0.000069 |
|  | Santa Cruz | 19.21 | 0.000016 |
| United States | New York | 5.33 | 0.000028 |
|  | Seattle | 6.40 | 0.000054 |
|  | Los Angeles | 16.38 | 0.000005 |

As a general feature of Table 2, we notice that

- In 11 out of the possible fifteen comparisons, the average cumulative rate of confirmed cases is higher when the average daily temperature is lower, and in the four remaining cases the differences in either variable are small.

A more specific analysis per country shows that

- In Iran, the average daily temperature in Bandar Abbas was the highest (21.54 °C), and the average cumulative rate of daily of confirmed cases was the lowest (0.000027).
- In Italy, the average daily temperature in Milan was the lowest (7.99 °C), and the average cumulative rate of daily of confirmed cases was the highest (0.000690).
- In Germany, the average daily air temperatures of the selected towns have similar values, as well as the spread of the COVID-19 infection is similar.
- In Spain, the average daily air temperature in Barcelona and Valencia are similar, and the spread of the infection is also similar. However, the comparison of Santa Cruz with higher average daily temperatures shows a slower spread of COVID-19.
- In the United States, the average daily air temperature in both New York and Seattle are lower than Los Angeles, and the average cumulative rate of confirmed cases shows the spread of the virus in both is faster than in Los Angeles.

These qualitative results reflect in the statistical analysis of the data by F-test and Independent-Samples T-Test presented in Table 3.

**Table 3.** Statistical analysis of the possible relation between the daily temperature and the average cumulative rate of confirmed cases, SPSS results

| Country | Case Study | Average Daily Temperature, °C |  |  |  | Average Cumulative Rate of Confirmed Cases |  |  |  |
| --- | --- | --- | --- | --- | --- | --- | --- | --- | --- |
|  |  | F-test |  | t-test |  | F-test |  | t-test |  |
|  |  | F Value | p-Value | t value | P-Value | F Value | P-Value | t value | P-Value |
| Iran | Ramsar | 20.4 | .000 | -1.93 | .048 | 16.33 | .000 | 2.697 | .012 |
|  | Tehran |  |  |  |  |  |  |  |  |
|  | Ramsar | 2.9 | .096 | -33.07 | .000 | 59.65 | .000 | 5.129 | .000 |
|  | Bandar Abbas |  |  |  |  |  |  |  |  |
|  | Tehran | 30.1 | .000 | -13.76 | .000 | 34.31 | .000 | 4.186 | .000 |
|  | Bandar Abbas |  |  |  |  |  |  |  |  |
| Italy | Milano | .12 | .731 | 6.63 | .000 | 50.16 | .000 | -4.720 | .000 |
|  | Rome |  |  |  |  |  |  |  |  |
|  | Milano | .95 | .333 | 8.84 | .000 | 52.49 | .000 | -4.778 | .000 |
|  | Naples |  |  |  |  |  |  |  |  |
|  | Rome | .90 | .346 | 2.21 | .054 | 2.54 | .119 | -.608 | .547 |
|  | Naples |  |  |  |  |  |  |  |  |
| Germany | Berlin | .00 | .998 | -1.83 | .071 | .020 | .887 | -.682 | .492 |
|  | Dusseldorf |  |  |  |  |  |  |  |  |
|  | Berlin | 2.23 | .140 | -.76 | .448 | .434 | .514 | .115 | .909 |
|  | Munich |  |  |  |  |  |  |  |  |
|  | Dusseldorf | 2.02 | .160 | .86 | .394 | .55 | .463 | .835 | .409 |
|  | Munich |  |  |  |  |  |  |  |  |
| Spain | Barcelona | 9.79 | .003 | 1.47 | .146 | .27 | .609 | .199 | .843 |
|  | Valencia |  |  |  |  |  |  |  |  |
|  | Barcelona | .019 | .890 | -16.44 | .000 | 13.48 | .001 | 2.183 | .048 |
|  | Santa Cruz |  |  |  |  |  |  |  |  |
|  | Valencia | 7.50 | .008 | 5 | .000 | 20.60 | .000 | 2.183 | .041 |
|  | Santa Cruz |  |  |  |  |  |  |  |  |
| United States | New York | 19.74 | .000 | -1.40 | .170 | 2.51 | .121 | -1.531 | .134 |
|  | Seattle |  |  |  |  |  |  |  |  |
|  | New York | 6.89 | .011 | -13.31 | .000 | 10.72 | .002 | 2.123 | .047 |
|  | Los Angeles |  |  |  |  |  |  |  |  |
|  | Seattle | 5.25 | .025 | -19.67 | .000 | 44.462 | .000 | 3.825 | .001 |
|  | Los Angeles |  |  |  |  |  |  |  |  |

In Table 3. t values or p-values can be used to check the existence of a significant correlation between the data for two cities.

Let us discuss in detail one such comparison, that between Ramsar and Tehran. From the F-test, the F value is equal to 20.4. The criterion for variance equality in the F-test is that the F value must smaller than the critical F value. The statistical analysis has been done on a statistical confidence level of 95%, and therefore the error level is 0.05, and considering the degrees of freedom, the critical F value of the test that can be found through F distribution tables [42], in this case, is equal to F = 1.57. Since the F value we found is 20.4, larger than the critical F value, the assumption of the equality of variances is rejected. It means that the average temperature variance of the Ramsar and Tehran are not equal.

According to the t-test, the t value for these two cities is equal to -1.929. Based on an error level of 0.05 and by considering the degrees of freedom, the critical values of the test indicated in the T distribution tables [43], is in this case equal to t = ± 1.645. Since the t value is out of the critical boundary (-1.645 ≤ acceptable t ≤ +1.645), the assumption of equality in the averages is rejected, and the existence of difference in averages is confirmed. Thus from two tests, it is evident that there is a significant difference between the average temperature of the two cities of Ramsar and Tehran.

An analogous result holds for the average cumulative rate of confirmed cases.

The main results of Table 4, with a confidence level of 95% are as follow:

- In Iran, the city of Bandar Abbas is significantly different from Ramsar and Tehran in terms of both average daily temperature and average cumulative rate of confirmed cases.
- In Italy, the city of Milan is significantly different from Rome and Naples in terms of both average daily temperature and average cumulative rate of confirmed cases. Instead, the two cities of Rome and Napoli are not significantly different in terms of these variables.
- In Germany, there are no significant differences in average daily temperature in all three cities, and the average cumulative rate of confirmed cases is not significantly different.
- In Spain, Barcelona and Valencia are not significantly different in terms of average daily temperature, and the average cumulative rate of confirmed cases is not significantly different. However, in Barcelona and Santa Cruz, the average daily temperature is significantly different, as well as the average cumulative rate of confirmed cases, and the same trend exists between Valencia and Santa Cruz.
- In the United States, the two cities of New York and Seattle are not significantly different in terms of average daily temperature, and of the average cumulative rate of confirmed cases, the number of patients is not significantly different. Instead, the city of Los Angeles is significantly different in terms of average daily temperature with two cities of New York and Seattle, and the average cumulative rate of confirmed cases are different significantly.

## 4. Discussion

The results of our analysis of daily air temperature for three case studies from five countries showed the existence of correlations between the average temperatures and the epidemic rate, and this is especially evident when the differences in average daily temperature are larger. For example, in Iran and Italy, the regions with lower average daily temperature had a higher epidemic level of the new coronavirus (COVID-19), and the same trend exists in all other case studies.

The statistical analysis showed that in all the case studies we considered, referring to five different countries, if there is a significant difference in daily mean temperature, there is also a significant difference in the average cumulative daily rate of confirmed cases, supporting the proposed research hypothesis about the correlation between air temperature and the epidemic rates of COVID-19.

The results of this study are in good agreement with other researches about the impact of air temperature on epidemic diseases and COVID-19, such as the role of daily weather parameters on the confirmed cases by Pirouz et al., 2020 or the negative impact of cold weather on the spread of flu virus by Price et al., 2019.

Due to the correlations among temperature and hours of sunshine for future research, it is recommended to study the effects of the sun’s UV rays as well as the average hours of sunshine on the epidemic rates of COVID-19.

One of the limitations of our work is due to the fact that in our analysis a possible correlation between the effects induced by temperature and those induced by other factors (e.g.social distancing, environmental pollution, seniority of the population, sanitation, lifestyles, etc.) was not taken into account.

## 5. Conclusions

This study analyzed the possible correlation of daily average air temperature with the epidemic rate of the coronavirus (COVID-19) using significant differences analysis between the datasets. The research hypothesis is that in regions with a significant difference in the average daily temperature, the average cumulative daily rate of confirmed cases would also is different significantly. Conversely, to the absence of a significant difference in daily temperature, would correspond no significant difference in the rate of confirmed cases. The hypothesis was investigated through statistical analysis. In order to minimize the impact on the results of factors like the policy of the government or cultural differences among countries (food, exercise, weight, etc.), three case studies within five countries, namely Iran, Italy, Germany, Spain, and United States were compared separately. This comparison using statistical analysis through F-test and Independent-Samples T-Test of the daily temperatures and daily rate of confirmed cases shows that there is a correlation between the average temperature and the epidemic rate, and this is especially evident when differences in average daily temperature are significantly larger, as it happens for Bandar Abbas in Iran, Milan in Italy, Santa Cruz in Spain, and Los Angeles in the US. Besides, the analysis of the average air temperatures shows that the epidemic rates of COVID-19 were higher in the case studies with a lower average temperature. Instead, when no significant differences exist in the average daily temperature of two cities in the same country, there is no significant difference in the average cumulative daily rate of confirmed cases. In conclusion, the results of this study support the research hypothesis and confirm the effectiveness of the proposed method for analysis of the epidemic rates.

## Data Availability

All weather data and confirmed cases of COVID-19 are available online

https://www.worldometers.info/coronavirus/

https://www.ogimet.com/ranking.phtml.en

## Acknowledgement

We are grateful to Professor Manlio Gaudioso for his critical reading of the manuscript.

## Funding

This research received no external funding.

## Conflicts of Interest

The authors declare no conflict of interest.

## Notes

### Competing Interest Statement

The authors have declared no competing interest.

